# Capturing clinical progression in multisystemic genetic ataxias: lessons from a prospective study of 884 patients with autosomal recessive or early-onset ataxia

**DOI:** 10.1101/2022.10.05.22280687

**Authors:** Andreas Traschütz, Astrid D. Adarmes-Gomez, Mathieu Anheim, Jonathan Baets, Bernard Brais, Cynthia Gagnon, Janina Gburek-Augustat, Sarah Doss, Hasmet A. Hanagasi, Christoph Kamm, Peter Klivenyi, Thomas Klockgether, Thomas Klopstock, Martina Minnerop, Alexander Münchau, Mathilde Renaud, Filippo M. Santorelli, Ludger Schöls, Andreas Thieme, Stefan Vielhaber, Bart P. van de Warrenburg, Ginevra Zanni, Ralf-Dieter Hilgers, PREPARE consortium, Matthis Synofzik

## Abstract

**Objective:** The Scale for the Assessment and Rating of Ataxia (SARA) is the most widely applied clinical outcome assessment (COA) for genetic ataxias, but presents metrological and regulatory challenges. To facilitate trial planning, we characterize its responsiveness (including subitem-level relations to ataxia severity and patient-focused outcomes) across a large number of ataxias, and provide first natural history data for several of them.

**Methods:** Subitem-level correlation- and distribution-based analysis of 1637 SARA assessments in 884 patients with autosomal-recessive/early-onset ataxia (370 with 2-8 longitudinal assessments), complemented by linear mixed-effects modeling to estimate progression and sample sizes.

**Results:** While SARA subitem responsiveness varied between ataxia severities, *gait*/*stance* showed a robust granular linear scaling across the broadest range (SARA<25). Responsiveness was diminished by incomplete sub-scale use at intermediate or upper levels, non-transitions (“static periods”), and fluctuating decreases/increases. All subitems -except *nose-finger*-showed moderate-to-strong correlations to activities of daily living, indicating that metric properties -not content validity-limit SARA responsiveness. SARA captured mild-to-moderate progression in many genotypes, e.g., SYNE1-ataxia: 0.55 points/year, AOA2: 1.14, POLG-ataxia: 1.56; but no change in others (ARSACS, COQ8A-ataxia). While sensitivity to change was optimal in mild ataxia (SARA≤10), it substantially deteriorated in advanced ataxia (SARA>25; 2.7-fold sample size). Use of a novel rank-optimized SARA without subitems *finger-chase* and *nose-finger* reduces sample sizes by 20-25%.

**Interpretation:** This study comprehensively characterizes COA properties and annualized changes of the SARA across and within a large number of ataxias. It suggests specific approaches for optimizing its responsiveness that might facilitate regulatory qualification and trial design.

## Introduction

With mechanistic treatment trials on the horizon for many genetic ataxias, sensitive capture of clinical treatment response has become key for academia, industry, and regulatory agencies. The Scale for the Assessment and Rating of Ataxia (SARA) serves as the most widely applied primary clinical outcome assessment (COA, specifically: clinician-reported outcome (ClinRO)) for almost all genetic ataxias^1, 2^. The SARA has been applied in large observational cohort studies to estimate disease progression and trial sizes in Spinocerebellar Ataxia (SCA) Type 1, 2, 3 and 6, or Friedreich Ataxia (FA)^3, 4^, and as primary endpoint in several randomized controlled trials^5-11^.

However, while validity and reliability of the SARA are excellent^1, 12-14^, regulatory agencies and recent studies in SCA3 have raised concerns given its insufficiently understood sensitivity to change and functional relevance, especially at the level of single subitems^15-17^. Repeated home-based video assessments have recently highlighted the strong intra-individual variability of the SARA^18^, which might be particularly driven by certain subitems. Although designed as a scale for a single construct of ataxia, the SARA score level is influenced by multisystemic features variably present across most ataxia genotypes (e.g., neuropathy, spasticity, or non-ataxia movement disorders), but their functional effect on the SARA properties are yet unclear. Together, these limitations of the SARA present a major challenge in current trial planning. While modifications to optimize the SARA are thus now being discussed (e.g. the modified SARA score *f-SARA*^*19*^), the underlying data evidence and validation of such modifications has remained scarce^16^. Extensive real-world data from ataxia registries, with prospective SARA assessments across many years and ataxia genotypes, may in turn provide the opportunity for statistical modeling and data-driven optimization of its responsiveness as a generic COA of ataxia.

Harnessing a large multi-center prospective cohort study of autosomal recessive and early-onset ataxias, we here provide an in-depth analysis of SARA’s ability to capture change, both across (i.e. as generic COA) and within (i.e. as genotype-specific COA) a large number of ataxias. Specifically, we (i) analyze its responsiveness at the total score as well as the subitem level; (ii) identify the influence of non-ataxia features and ataxia severity; (iii) characterize the relation to patient-focused outcomes; and (iv) simulate data-driven rank-optimized SARA composites for trial size estimations. In addition, we (v) provide first prospective natural history data for a number of genetic ataxias to facilitate the design of treatment trials.

## Methods

### Study cohort

Prospective cross-sectional and longitudinal data from all 948 consecutive patients enrolled between 2013 and August 4th 2021 were retrieved from the global multicenter ARCA Registry^20^. Datasets included (i) genotypic and demographic data, (ii) assessments of ataxia severity (SARA^1^) and non-ataxia features (including the Inventory of Non-Ataxia Signs (INAS)^21^), and (iii) the functional staging (FARS-FS) and activities of daily living (FARS-ADL) scales of the Friedreich Ataxia Rating Scale (FARS)^22^ as patient-focused outcomes. Patients had been eligible for inclusion into the ARCA Registry if they had (i) a genetically confirmed autosomal recessive cerebellar ataxia (ARCA), and/or (ii) an early-onset ataxia (EOA) with onset before age 40 years without evidence of an autosomal dominant family history, repeat-expansion SCA or acquired cause (e.g., subacute onset, rapid progression, alcohol intake, abnormal B12 levels, CSF pleocytosis, or structural lesions on imaging), thus representing a stratum of ataxia patients known to be enriched for ARCAs^23, 24^. Discarding eligibility failures and datasets with neither SARA nor phenotypic data, a total of 931 ARCA/EOA patients were included in the final analysis. A subset of nine ARCAs recurring in at least 15 patients was grouped as ‘common ARCAs’ (n=393 patients) to enable statistical analyses adjusting for genotype. Datasets included overall 1637 SARA assessments (n=884 patients; 370 with 2-8 longitudinal assessments; 86 genotypes), at least one INAS assessment in 908 patients, 187 assessments with FARS-FS, and 62 assessments with the FARS-ADL. At each contributing site, patients had provided informed consent for pseudonymized data entry in the ARCA Registry.

### Statistical analysis

#### Prevalence and influence of non-ataxia features

The presence of multisystemic non-ataxia features was determined from each patient’s last available INAS and the registry’s systematic history for *a priori* recurrent features of ARCAs (e.g., epilepsy or mitochondrial features such as hearing loss or diabetes; case report form “Clinical Features”^20^). Their respective impact on the SARA as a measure of ataxia severity was analyzed by generalized linear modeling (GLM), using the last SARA score of each patient as dependent variable; selecting age of onset, ataxia duration, ataxia duration squared, and genotype as independent variables^25^; and adding non-ataxia features in the corresponding assessment that occurred in at least 40 patients (i.e. >∼5% prevalence) individually as independent factors to the model. To account for multiple comparisons of non-ataxia features, *p*-values were adjusted by the Benjamini-Hochberg method with a false discovery rate of 0.05. Only non-ataxia features with independent replication in separate GLMs including the most common (genetically solved and stratified) ARCAs and all unsolved EOAs, respectively, were considered significant generic determinants of ataxia severity.

#### Correlation-based analysis of SARA subitems

The responsiveness (here defined as intra-item evolution along disease severity) of SARA subitems across the full range of ataxia severity was examined by correlating each subitem score to the corresponding total SARA score, and by fitting third order polynomial curves to estimate response curves and 95% prediction intervals^26^. To analyze the homogeneity of subitem response curves across ataxias, this analysis was performed across all 1637 assessments in the cohort, and independently for each common ARCA as compared to all other ARCAs/EOAs. Patient-focused functional relevance was determined by correlating the SARA and its subitems to the FARS-ADL, a 9-item composite scale of basic activities of daily living (speech, cutting food/handling utensils, dressing, personal hygiene, falling, walking, sitting) and impairments (swallowing, bladder function).

#### Progression analysis

Annualized progression rates for the SARA and its subitems were estimated using LMEM (REML, covariance structure: variance components, Kenward-Roger degrees of freedom) with random effects on intercept (score at baseline) and slope (points per year of follow-up)^4^. Annualized progression rates were compared between ataxia severities, SARA subitems, and different composites by adding interaction terms between follow-up time (months since baseline visit/12) and the respective groups. Ataxia severity was binned according to the SARA at baseline (≤10, 10-25, and >25 for mild, moderate, and advanced ataxia), and separated into ambulatory and non-ambulatory patients (operationalized by SARA subitem *gait* >6, indicating inability to walk more than 10 meters even with strong support or full inability to walk). While always arbitrary, the pragmatic binning according to baseline SARA was data-driven, aiming to capture relevant floor, ceiling, and plateau segments of SARA subitems in cross-sectional data (**Fig. 2**). All bins were evaluated by comparing available FARS-FS levels as external anchor. Genotype (of the common ARCA genotypes) was added as fixed effect to the LMEM in control analyses, allowing to capture both genotype-specific effects but also across-genotype effects on disease progression. We considered for potential effect of ceiled and floored observation on the model fit and compared our results for LMEM with Tobit models. To compare sensitivity to change between different SARA composites and ataxia severities, we calculated hypothetical sample sizes (1:1 allocation ratio) based on the progression estimates from the LMEM, simulating a 50% reduction in SARA progression in an arbitrary parallel group, 2-year, 5-visit interventional trial with α=0.05 and 90% power.

Annualized progression rates (mean, standard error (SE), and 95% confidence intervals (CI)) and sample sizes were calculated with SAS, version 9.4 (procedure MIXED, NLMIXED). SPSS 25 (IBM Corp, Armonk, NY) was used to perform GLMs (function GENLIN). Correlations, curve fitting, descriptive, and confirmatory statistics were calculated with GraphPad Prism 9 (GraphPad Software, La Jolla, CA), using non-parametric measures (Spearman’s rho, median, interquartile range (IQR)). MATLAB 2019b (The MathWorks, Natick, MA) was used to determine bootstrapped 95% CIs of Spearman correlations.

## Results

### Baseline characteristics

We included 931 patients (51% female, median age at baseline: 36 years, range: 2-83, **Fig. 1A**) with genetically confirmed autosomal recessive cerebellar ataxia (ARCA) or genetically unsolved early-onset ataxia (EOA) highly enriched for ARCA of global origin, predominantly from Europe (n=645), followed by Canada (n=109), Asia (n=96), and Africa (n=47). The genetic cause of ataxia was identified in 623/931 patients (67%, 86 genotypes; see **Fig. 1A**,**B** for frequency of ‘common ARCAs’). Disease onset in the genetically confirmed ARCA cases was before age 40 years in 94%, thus supporting the pragmatic cut-off of age of onset <40 years chosen for the genetically still unsolved cohort. The cohort spanned the full range of ataxia severities (SARA from 3-40, **Fig. 1B**) across the common ARCAs, thus allowing for detailed metrological SARA analyses, including disease progression analysis.

**Figure 1:**
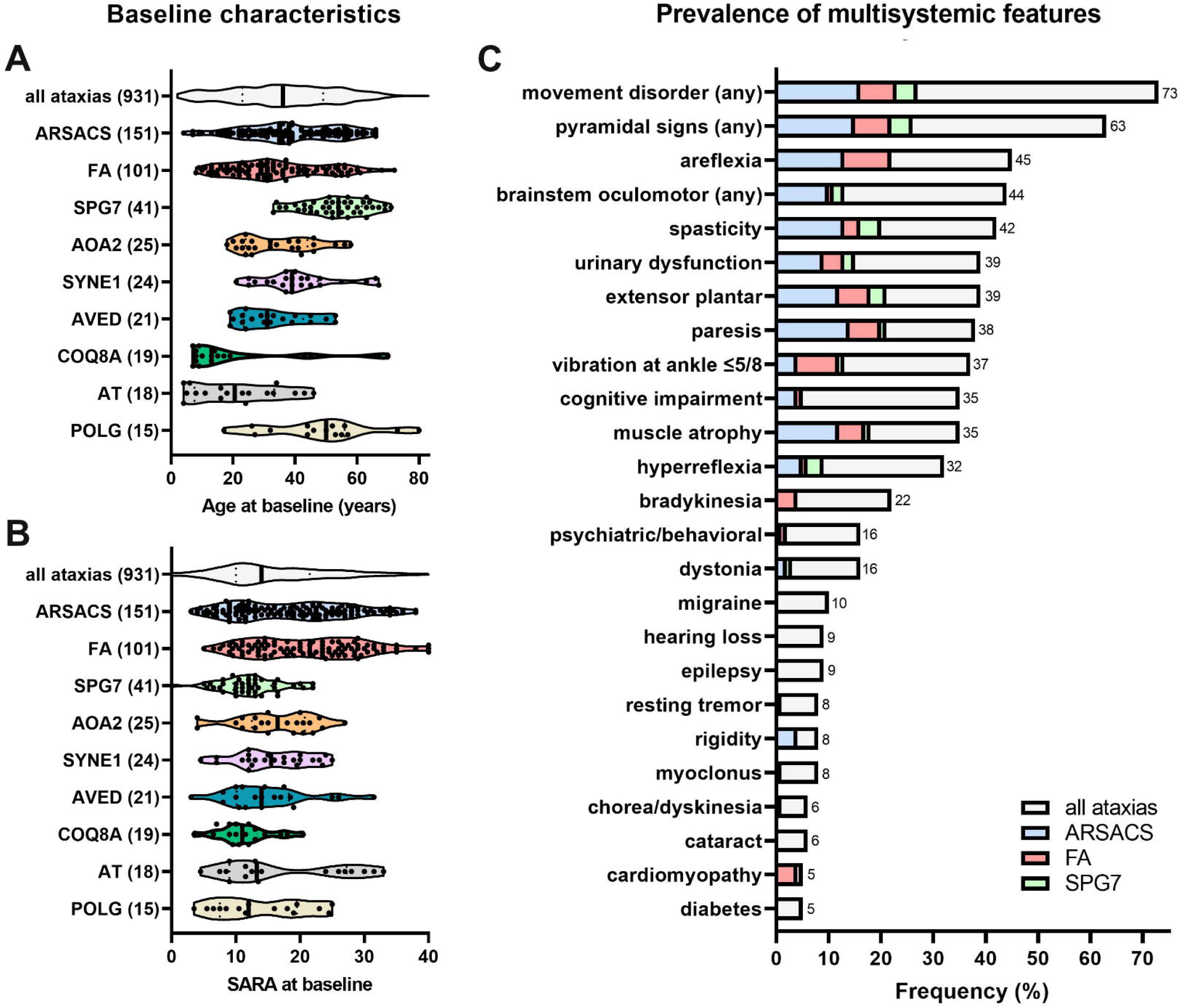
Characterization of multisystemic ataxia cohort. Characterization of patient age (A) and ataxia severity (B) at baseline assessment for the autosomal recessive and unsolved early-onset cerebellar ataxias that are part of this study. Patients in this study represent a wide distribution of these features, particularly also in the ‘common ARCAs’ (color-coded). Solid and dashed lines in violin plots indicate median and quartiles. Number of patients shown in brackets. (C) Frequency of extra-cerebellar features observed in at least 5% of patients. Multisystemic phenotypes were representative for the ataxia cohort, and -with the exception of cardiomyopathy-not predominantly caused by the three most common genotypes. Movement disorder (any) = any movement disorder other than ataxia. AOA2 = Ataxia with oculomotor apraxia type 2; ARSACS = Autosomal recessive spastic ataxia of Charlevoix-Saguenay; AT = Ataxia-teleangiectasia; AVED = Ataxia with vitamin E deficiency; FA = Friedreich ataxia; SPG7 = Spastic paraplegia type 7; COQ8A/POLG/SYNE1 = ataxia related to respective gene.

**Figure 2:**
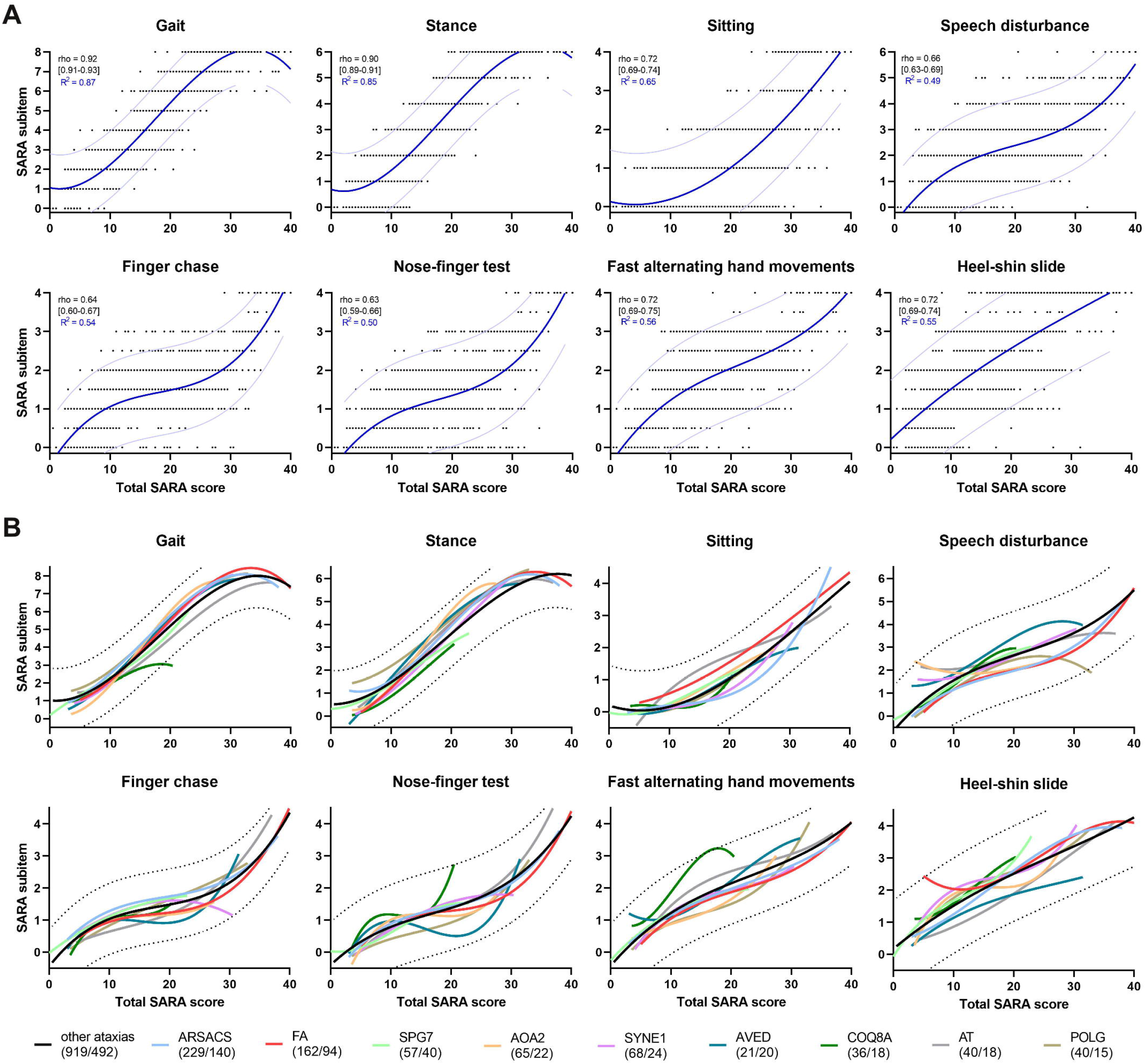
Responsiveness of SARA subitems across ataxia severity and genotypes. Cross-sectional analysis of 1637 SARA assessments in 884 patients. (A) Spearman correlation [95%CI] between SARA subitem and total SARA (=overall ataxia severity). Response curves and 95% prediction intervals estimated by third order polynomial curve fits. SARA subitems show ceiling effects (*gait, stance*), floor effects (*sitting*), and non-responsive “static” periods at moderate ataxia severity (particularly *speech, finger chase* and *nose-finger*). (B) Comparison of polynomial response curves between the ‘common ARCAs’ against all other ataxias in the cohort. Numbers in brackets indicate total number of SARA assessments per number of patients. The relative contribution of each subitem to the total SARA is overall similar across ataxias, with genotype-specific differences particularly for COQ8-ataxia.

### Non-ataxia features determine ataxia severity

ARCAs and EOAs were inherently multisystemic, with an INAS count of zero (indicating absence of non-cerebellar involvement) in only 4% of patients (median INAS count: 5, IQR: 3-6). Non-ataxia movement disorders (73%; bradykinesia, rigidity, resting tremor, dystonia, myoclonus, and/or chorea/dyskinesia), pyramidal signs (63%; spasticity, extensor plantar response and/or hyperreflexia), and signs of neuropathy (areflexia: 45%) were particularly prevalent non-ataxia features (see **Fig. 1C** for complete list). Cognitive impairment was present in 35%, with predominant childhood onset (median: 6 years, IQR 2-24), and reportedly non-progressive in 59%. Ataxia severity as assessed by the SARA was independently associated with several non-ataxia features in both common ARCAs (n=393, adjusted for genotype) and unsolved EOAs (n=335). For example, SARA scores were up to 4-5 points higher if paresis was present or if vibration sense impaired in patients with otherwise similar genotype, age of onset, and disease duration (**Table 1**),

**Table 1:** Non-ataxia features associated with ataxia severity.

|  | Common ARCAs (n=393) |  |  | Unsolved EOAs (n=335) |  |  |
| --- | --- | --- | --- | --- | --- | --- |
| Feature | Beta | SE | p<sub>adj</sub> | Beta | SE | p<sub>adj</sub> |
| Paresis | 4.9912 | 0.8627 | <0.0001 | 4.8052 | 0.9916 | <0.0001 |
| Cognitive impairment | 4.4418 | 0.9927 | <0.0001 | 2.9897 | 0.9058 | 0.0023 |
| Vibration at ankle $\leq 5/8$ | 3.6171 | 0.8125 | <0.0001 | 3.9282 | 1.0253 | 0.0006 |
| Urinary dysfunction | 3.0997 | 0.7897 | 0.0006 | 5.4036 | 0.8950 | <0.0001 |
| Brainstem oculomotor sign (any) | 2.6768 | 0.8190 | 0.0039 | 3.8300 | 0.8951 | <0.0001 |
| Muscle atrophy | 2.6501 | 0.8135 | 0.0046 | 4.3656 | 1.0907 | 0.0005 |
| Spasticity | 2.1950 | 1.0034 | 0.0598 | 3.4616 | 0.9204 | 0.0008 |

Generalized linear model with SARA score as dependent variable, and genotype, age of onset, ataxia duration, and ataxia duration squared as independent variables. Non-ataxia features with a prevalence above 5% were individually entered into the model as independent factor. Listed features were identified in both common genetically defined ARCAs as well as in a separate cohort of unsolved EOAs. Beta = Regression coefficient, SE = standard error, p<sub>adj</sub> = p values adjusted by Benjamini-Hochberg method.

### Responsiveness of SARA subitems depends on ataxia severity

Individual subitems of the SARA showed variable responsiveness across the ataxia severity range (**Fig. 2A**). *Gait* and *stance* covered a large range of severities (87% SARA scores <25) with a linear scaling, including relatively granular progression steps of 8 and 6 metric levels, respectively, and relatively small, uniform prediction intervals even across the heterogeneity of ARCAs and EOAs in the cohort. In contrast, subitem *sitting* appeared only responsive for SARA scores above 10, but continued to increase with a linear slope until most advanced ataxia (SARA>30), where *gait* and *balance* contributed no longer to responsiveness. For subitems *speech, finger chase*, and *nose-finger*, responsiveness relative to the total SARA flattened in the range in which most patients were assessed (57% SARA scores 10-30), indicating “static periods” in an important SARA range, with steep increases only beyond that range. Accordingly, these subitems only showed moderate correlations with the total SARA score (rho: 0.6-0.7), as compared to strong correlations of all other subitems (rho: >0.7-0.9). In contrast, *fast alternating hand movements* captured ataxia severity also within the range of SARA 10-30, rendering this the most promising of all SARA upper limb subitems to capture appendicular upper limb ataxia. For appendicular lower limb ataxia, *heel-shin slide* was responsive across the full range of SARA scores, although with higher variability as compared to truncal ataxia items (*gait, stance, sitting*).

Despite the heterogeneity of ARCAs (including partly very different neural systems’ damage, for example paresis, spasticity, or sensory neuropathy), independent polynomial fits for each of the common ARCA genotypes as compared to all residual ataxias (non-common ARCAs and unsolved EOAs) revealed a relatively uniform progression pattern for each subitem relative to the total SARA, particularly seen for *gait* and *stance* (**Fig 2B**). A notable exception was COQ8-ataxia: here the contribution of individual subitems to the total SARA was lower for item *gait*, but higher for items *nose-finger* and *alternating hand movements* items (both compared to the 95% prediction interval of other ataxias.) This might be due to the highly prevalent upper limb movement disorders in COQ8A-ataxia such as tremor, myoclonus and dystonia.

### Metric limitations of SARA responsiveness

The analysis of the distribution of SARA subitem scores across all 1637 assessments (**Fig. 3A**), and of the longitudinal score level changes of each subitem across all 202 patients with a follow-up assessment after 1 year (mean: 12.8 months, range: 9-19) -as a proxy for a standard trial duration-(Sankey diagram, **Fig. 3B)** showed several metric limitations for the SARA responsiveness. Incomplete coverage of the full range of the scale was observed for all subitems. Intermediate score levels were under-used in subitems *gait* (5 = “Severe staggering, permanent support of one stick or light support by one arm required”), *stance* (4 = “Able to stand for >10 s in natural position only with intermittent support”), and *sitting* (3 = “Able to sit for > 10 s only with intermittent support”). Upper score levels were under-used in subitems *speech, finger-chase, nose-finger, and alternating hand movements*, where less than 1-6% of patients were rated with the highest score. Inconsistent transitions between score levels comprised: a substantial share of subitems with (neurologically unexpected) score decreases (“improvements”) from baseline to follow-up (particularly for *speech*); fluctuating large changes of > 1-2 points decreases (“improvements”) or increases (“deteriorations”) for subitems *finger chase* and *nose-finger*; and non-transition between score levels, for example from *speech* score 3 to 4 (“occasional” to “many words difficult to understand”).

**Figure 3:**
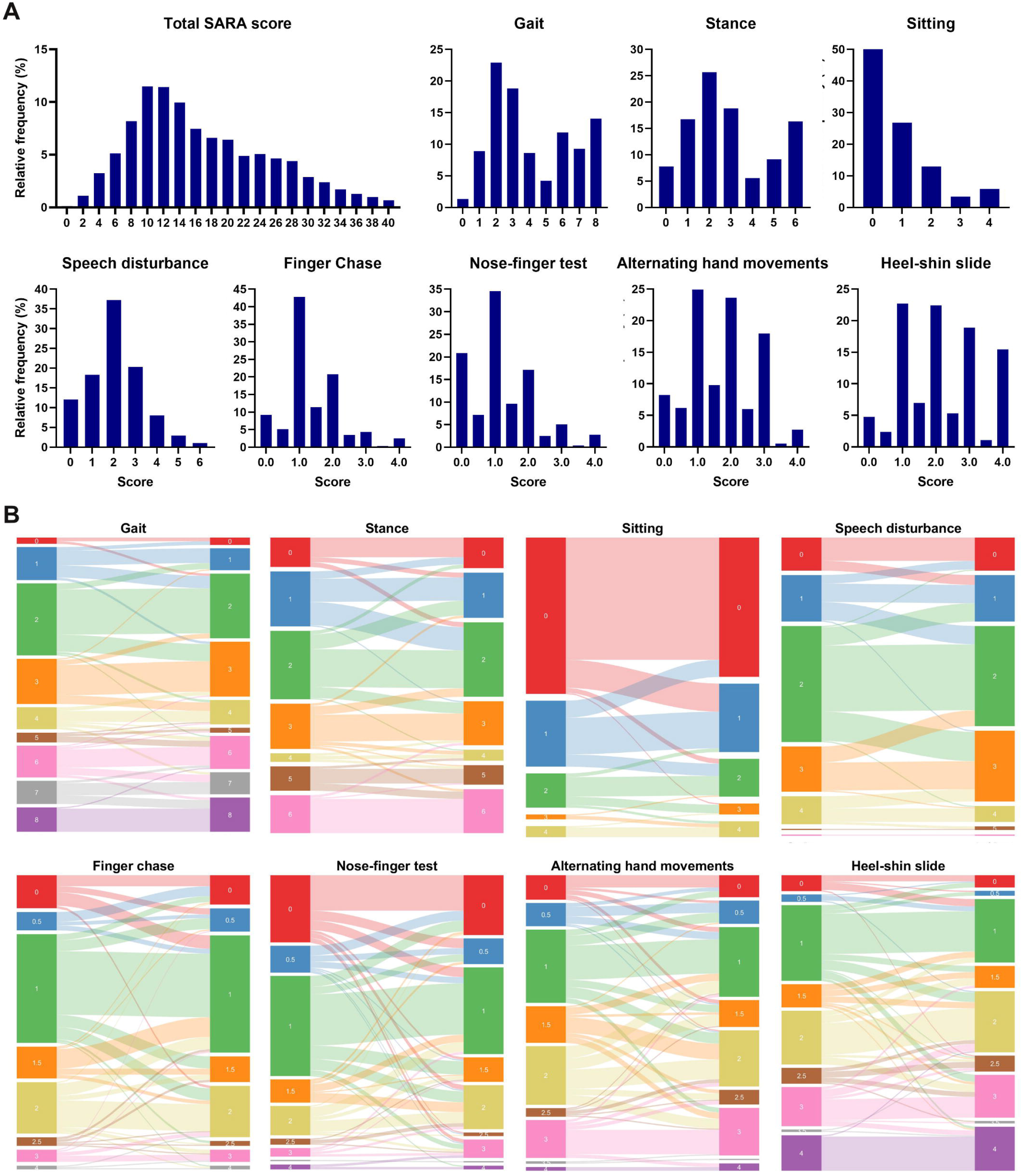
Distribution of the SARA subitems cross-sectional and in longitudinal progression. (A) Distribution of the SARA and its subitems across all 1637 assessments. While the total SARA score is widely distributed across the full score range, individual subitems show considerable ceiling effects (*gait, stance*) and floor effects (*sitting*), as well as under-use of intermediate scores (*gait, stance, and sitting*, but also uneven scores for appendicular items) and high scores (*speech, finger-chase, nose-finger*). (B) Sankey diagram of 202 patients with approximately annual follow-up. In addition to the incomplete use of score ranges seen in (A), SARA subitems with non-responsive periods were characterized longitudinally by substantial score decrease (“improvement”) for *speech* (scores 2 to 1, 3 to 2, and 4 to 3), and by volatile score decrease (“improvement”) and increase (“deterioration”) in appendicular items (e.g., scores 0 to 2 in finger chase and nose-finger, or scores 2 to 1 in all upper limb items.

### SARA correlations with patient-focused outcome

We next analyzed whether the limitations observed in the metrics of the SARA might be paralleled by (and thus possibly even be rooted in) limitations in its content validity, namely in reflecting differences in patient-focused outcomes. To this end, we correlated the SARA and its subitems to the FARS-ADL as a patient-focused anchor. The total SARA showed a strong correlation with FARS-ADL (rho: 0.85), indicating the capacity of this COA to capture everyday-relevant functional impairment across domains (**Fig. 4**). Strong correlations were also observed for its subitems *sitting* and *fast alternating hand movements* (rho: >0.7, comparable to subitems *gait* and *stance)*, and moderate correlations were still found for subitems *speech, finger chase*, and *heel-shin slide* (rho: >0.5). The functional relevance of these items thus indicates that they should not be discarded from the SARA, but rather need improvement in task design and/or scoring to increase their metric properties. In contrast, the weak correlation of *nose-finger* with FARS-ADL -thus paralleling its metric limitations-suggest that this SARA subitem might be of overall limited added value for the SARA as a COA of ataxia.

**Figure 4:**
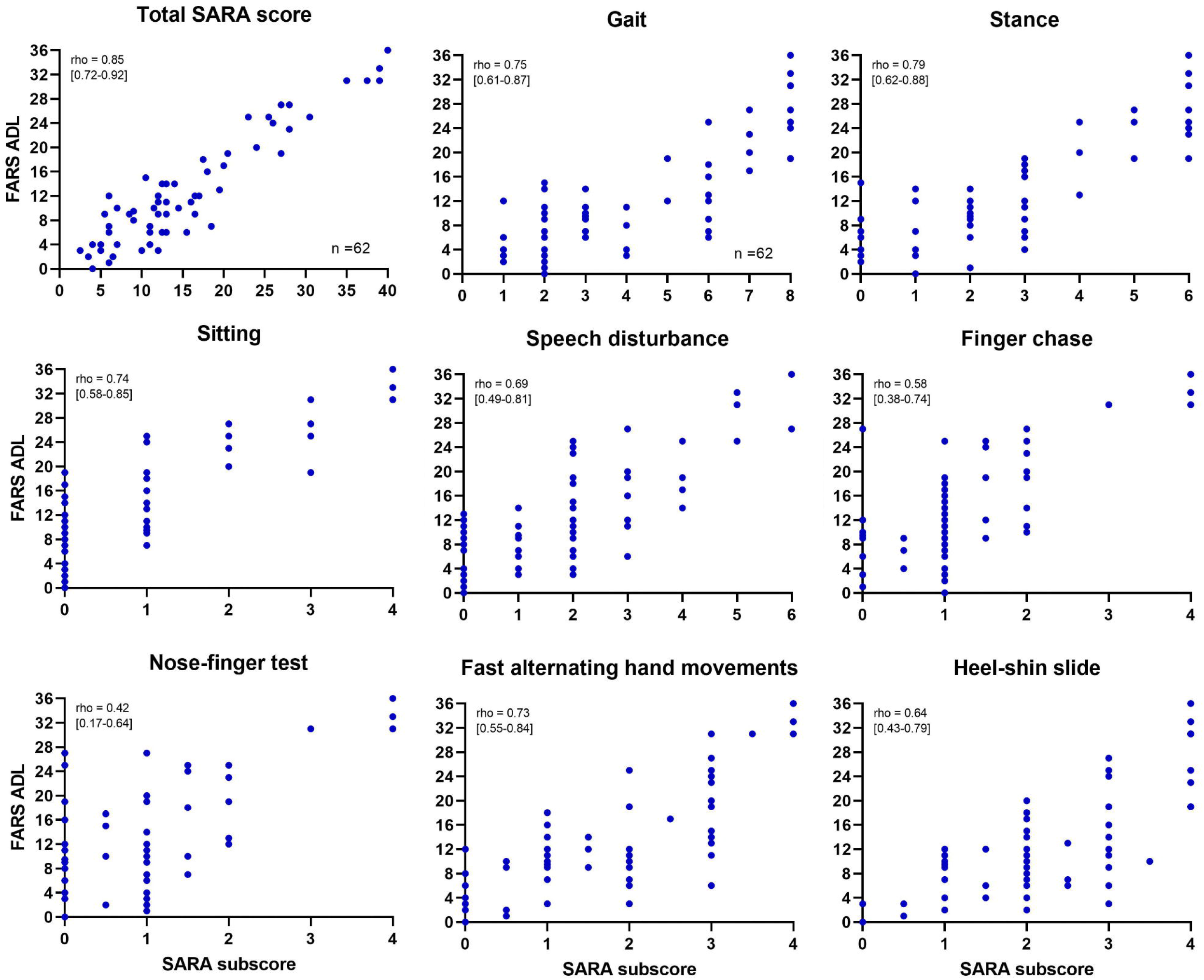
Correlation of the total SARA and its subitems with FARS-ADL. Spearman correlations with [95%CI]. The total SARA and all SARA subitems - except *nose-finger -* show at least moderate correlations with the FARS-ADL, which can be considered as a clinically meaningful endpoint.

### Modelling of longitudinal progression

The SARA was generally able to capture progression of ataxia severity in the cohort, with overall mild-to-moderate annualized progression across all ARCAs/EOAs (LMEM, +0.60 [SE: 0.06] points/year), but with substantial variability between genotypes, ataxia severities, and SARA subitems.

#### (i) Disease progression according to genotypes

Annual SARA progression was highest in POLG-ataxia (1.56 [0.32]), FA (1.49 [0.19]), and Ataxia with oculomotor apraxia type 2 (AOA2; 1.14 [0.24]). In contrast, SARA progression was only moderate in Spastic paraplegia type 7 (SPG7; 0.57 [0.41]), SYNE1-ataxia (0.55 [0.26]), and Ataxia-Teleangiectasia (AT; 0.62 [0.38]). No change within 1 year was captured by the SARA in Autosomal recessive spastic ataxia of Charlevoix-Saguenay (ARSACS; 0.12 [0.15]) and COQ8A-ataxia (−0.06 [0.41]; **Figure 5**).

**Figure 5:**
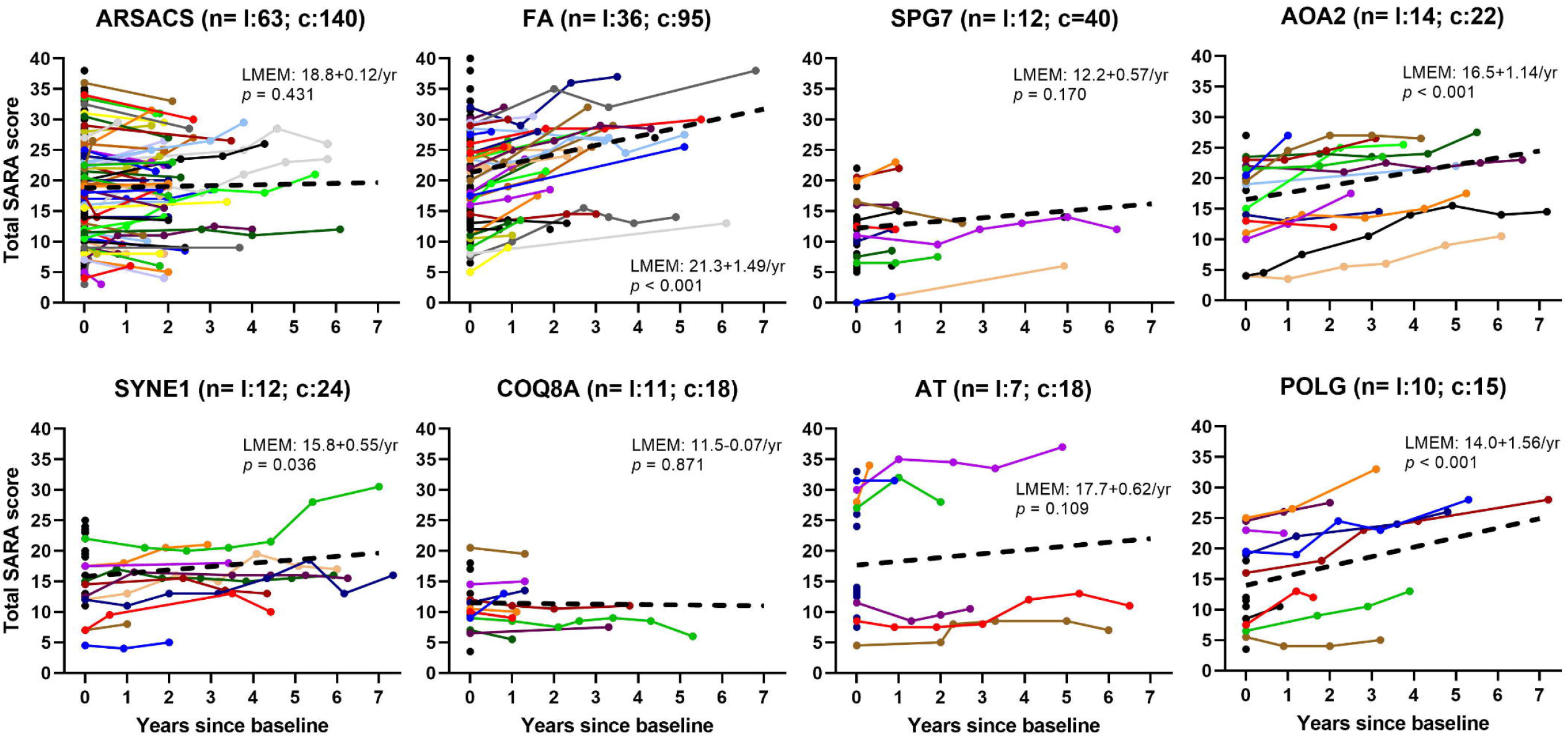
Annualized progression of the SARA in the most common ARCAs. Dashed lines and corresponding equations present LMEMs with random intercept and slope; *p* values <0.05 indicate progression significantly different from zero. l: number of subjects with *l*ongitudinal data (colored lines), c: total number of subjects including *c*ross-sectional baseline data without follow-up (black dots). The SARA captures change in FA, AOA2, SYNE1 und POLG, but does not show change in ARSACS or COQ8A-ataxia within 1 year. Intercepts of baseline SARA scores can be interpreted as expectable distributions of patients’ ataxia severity for trial planning. AVED not shown because only one patient had longitudinal data.

#### (ii) Disease progression according to ataxia severity

Ataxia severity was binned by baseline SARA (mild: <10, moderate: 10-25, advanced: >25) and by ambulatory status as critical ataxia milestone (baseline SARA gait >6 as proxy for loss of ambulation). This pragmatic binning was supported by the FARS-FS, which was available for a subset of assessments, and which differed between mild (n= 58, median: 2, [IQR: 1.5-2.5]), moderate (n=101, 3 [2-4]), and severe ataxia (n=27, 5 [4.5-6]; Kruskal-Wallis test with Dunn’s multiple comparisons: all *p*<0.001), as well as between ambulatory (n=149, 2.5 [2-3.5]) and non-ambulatory bins (n=38, 5 [5-5.1], Mann-Whitney test: p<0.001). Sensitivity to change of the SARA was similar in mild (0.67 [0.12]) and moderate ataxia (0.64 [0.09]), while progression decreased and variability increased in advanced ataxia 0.37 [0.19]; **Figure 6A**). Accordingly, progression decreased, and variability increased when ataxia severity was stratified by loss of ambulation (baseline SARA gait >6: 0.34 [0.18] vs. SARA gait ≤6: 0.63 [0.07]).

**Figure 6:**
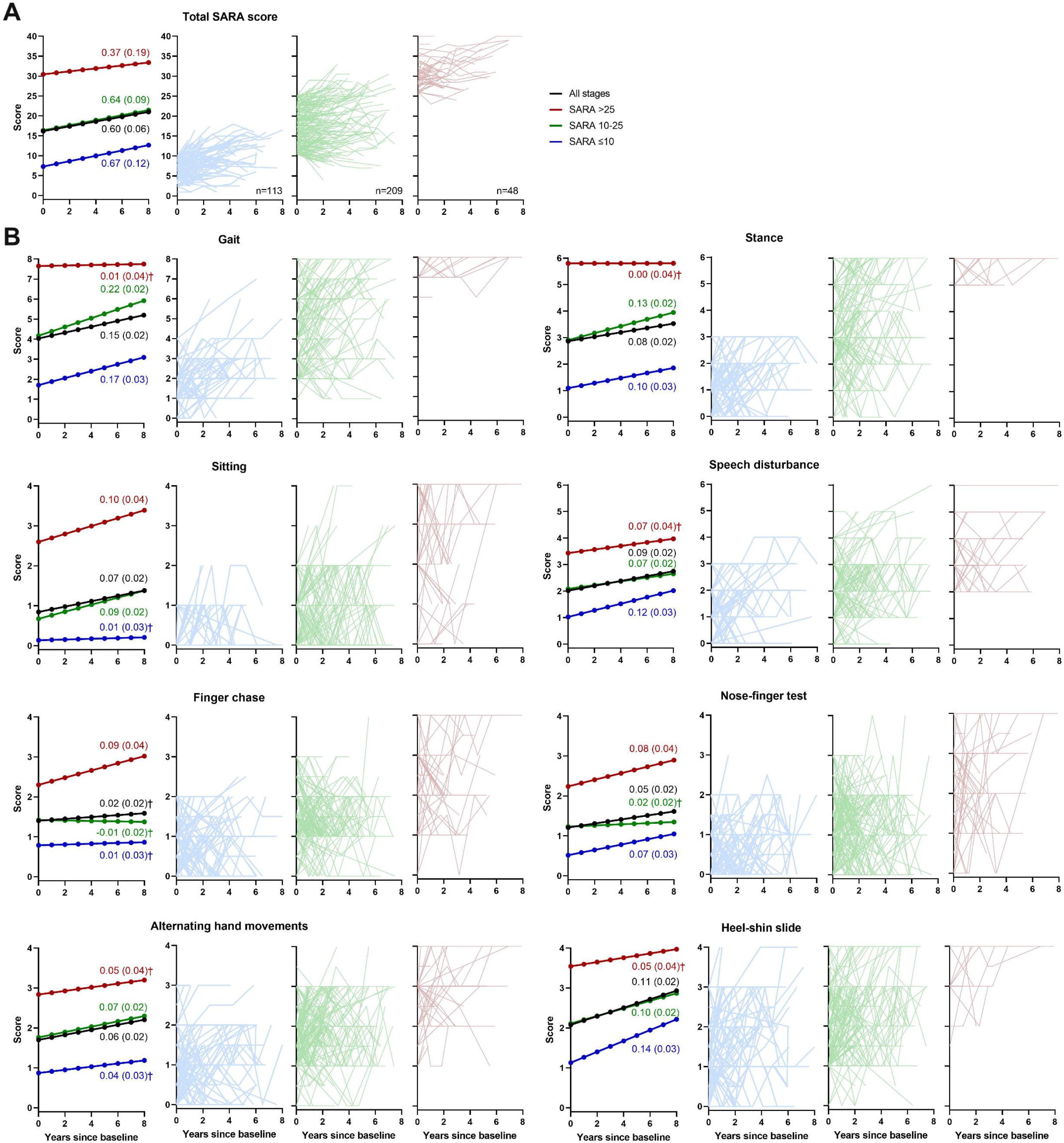
Sensitivity to change of the total SARA and its subitems by ataxia severity. Raw data and mean (standard error) estimated annualized progression based on linear mixed-effects modeling of all longitudinal data in the full cohort. Results are shown aggregated across all disease stages (black) as well as separate for the mild (SARA <10, blue), moderate (SARA 10-25, green), and advanced (SARA> 25, red) ataxia. Crosses (†) mark models without sensitivity to change. (A) Sensitivity of the SARA decreases in advanced ataxia due to smaller progression and higher variability. (B) At the level of SARA subitems, *gait, stance, speech*, and *heel-shin* show sensitivity to change in mild and moderate ataxia (i.e. SARA ≤25), thus likely driving the sensitivity to change of the total SARA in these disease stages. In advanced ataxia (i.e. SARA >25), *sitting, finger chase* and *nose-finger* show sensitivity to change, thus here likely driving the changes in the total SARA. Note the marked variability in the trajectory of individual patients.

#### (iii) Disease progression of SARA subitems

Overall, all SARA subitems - except *finger-chase-* were sensitive to change, with the largest annual progression for *gait*, followed by *heel-shin, speech*, and *stance* (**Fig. 6B**). All of these four most responsive items consistently lost sensitivity to change in advanced ataxia (SARA>25), while subitem *sitting* was not sensitive to change in mild ataxia (SARA<10; **Fig. 6B**). Among upper limb subitems, *alternating hand movements* was only sensitive to change in moderate ataxia, while *finger-chase* and *nose-finger* were sensitive in advanced ataxia despite their metric limitations.

### Rank-optimization of SARA for clinical trials

Based on the annualized progression estimated by LMEM, we performed hypothetical sample size calculations as a methodological tool to test and illustrate the sensitivity of different SARA composites in trial scenarios (rather than for use in actual trials, which will likely be conducted in a disease-/genotype specific fashion, except for symptomatic ataxia drugs). As use cases, we analyzed the sensitivity of the SARA across the full cohort and the subset of common ARCAs, which allowed to characterize and optimize its sensitivity as an overall, generic COA of ataxia (rather than for a particular genotype). Compared to a trial in ARCA/EOA patients with mild ataxia (SARA: ≤10, N=486), sample size increased 1.7-fold (N=808) when applying the SARA as COA in moderate ataxia (SARA: 10-25), and 2.7-fold (N=1306) in advanced ataxia (SARA: >25; **Fig. 7A**). In the common ARCAs, estimated sample sizes were similar in early and moderate ataxia (both N=440), but increased even 3.6-fold in advanced ataxia (N=1602).

**Figure 7:**
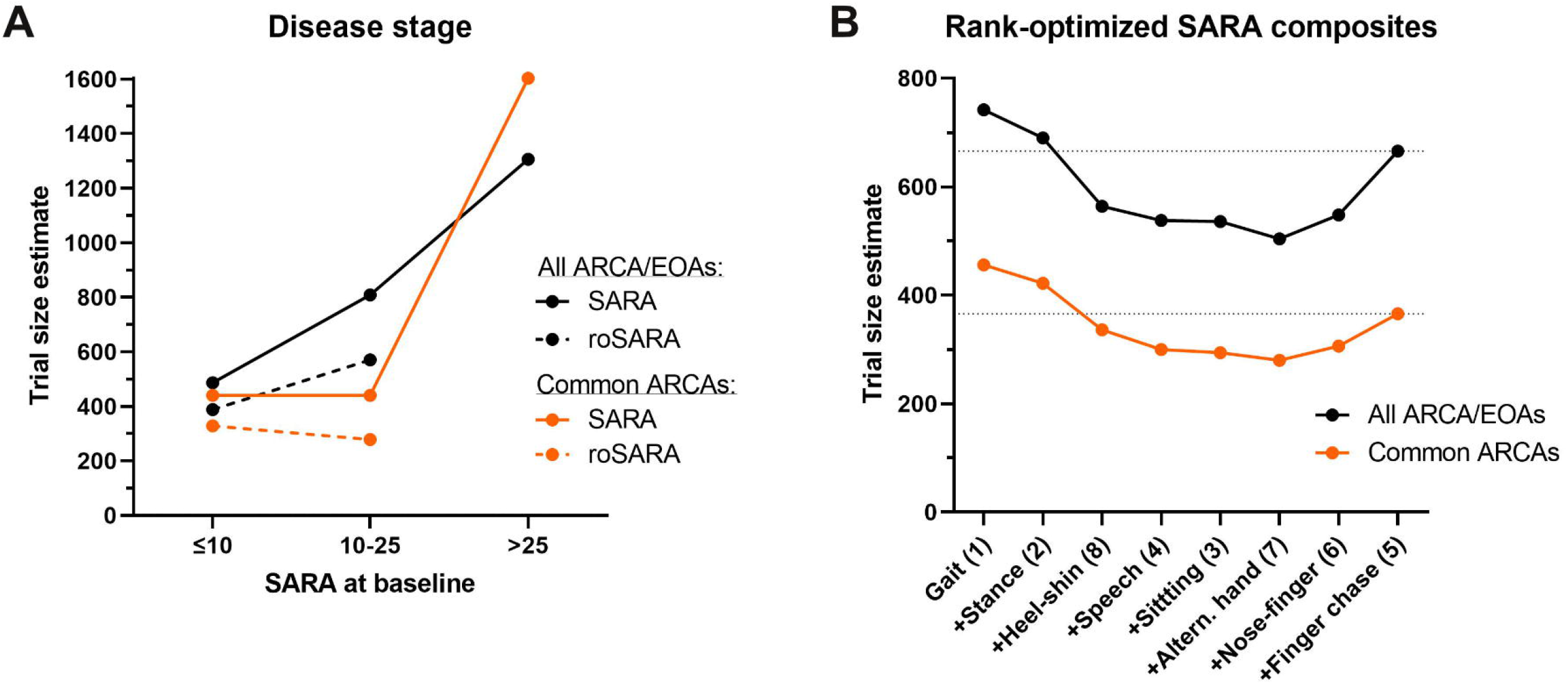
Sample size estimates for the SARA and a rank-optimized SARA (roSARA) relative to ataxia severity (A) and for rank-optimized SARA composites (B). Simulations illustrate the relative impact of altered sensitivity on hypothetical sample sizes in trial scenarios, estimated for the detection of 50% reduced progression in a 2-year 5-visit trial and a power of 90%. (A) Sample size estimates for the SARA depend on ataxia severity, with sharp increases (about 3fold) in advanced ataxia. The roSARA (dashed line) shows an improved sensitivity compared to the classical SARA (uninterrupted line), both for the full cohort (= all ARCAs/EOAs; black) and for the subgroup of the most common ARCAs stratified for genotype (orange). Sample size for roSARA was calculated only for the trial-relevant mild (SARA <10) and moderate (SARA 10-25) disease stage; data was insufficient for modeling the advanced stage. (B) Sensitivity of a series of rank-optimized SARA composites with successive step-by-step inclusion from the most (left side: *gait*) to least sensitive subitem (right side: *finger-chase*), calculated for baseline SARA≤25 points. Trial sizes would decrease by 20-30% if subitems *nose-finger* and *finger-chase* were omitted. Note the comparable sample sizes between the total SARA (dashed lines) and a composite of only 2-3 most optimal subitems.

We next explored whether sensitivity of the SARA could be optimized by including (and omitting) subitems according to their ranked annual progression rate, here focusing on the most trial-relevant severity range of ataxia, namely mild-to-moderate ataxia. Successive inclusion of the six most responsive subitems (*gait, stance, heel-shin, speech, sitting, alternating hand movements*) into such a generic rank-optimized SARA (roSARA), calculated across all ARCA/EOA patients with a baseline SARA≤25, led to a continuous decrease in sample size from N=742 to N=504, i.e. yielding a reduction of trial size by 18% for the roSARA as compared to total SARA (**Fig. 7B**). Adding *nose-finger* and *finger chase* to the roSARA, i.e. simulating all items of the SARA (=total SARA), worsened sensitivity (N=666), with a suboptimal level comparable to the sensitivity of only the 2-3 most responsive items. Similar results were obtained for the roSARA in the common ARCAs with a baseline SARA≤25, with a reduction of trial size by 24%.

## Discussion

To facilitate trial planning and regulatory qualification, this study comprehensively characterizes responsiveness of the SARA as generic COA for ataxia, harnessing a large prospective genetic ataxia cohort as exemplary show-case cohort for multisystemic ataxias, and provides first natural history data for a wide range of ataxias.

### Responsiveness of the SARA

As a major finding, our subitem-level metric analysis delineated three types of limitations that hamper responsiveness of the SARA. First, our correlation-based analysis showed limited responsiveness in several subitems and disease stages, with varying patterns: ceiling effects for *gait* and *stance*, floor effects for *sitting*, and non-responsive (‘static’) periods at moderate stages particularly for *speech, finger-chase*, and *nose-finger*. These findings extend partly similar recent observations in a monocentric FA^26^ and multicentric SCA3 cohort^15^, but now demonstrate these SARA properties for a more comprehensive set of items in a multicenter cohort and in particular across a large number of genetic ataxias, i.e. for the SARA as a *generic* COA *across* ataxias. Second, our distribution-based analysis identified incomplete coverage (“use”) of the full scale range in almost all subitems. Under-use of SARA *gait* score 5 may reflect the patients’ need for better stabilization by bilateral walking aids or a stroller once they lose free ambulation^27^. For *stance* and *sitting*, we hypothesize that “intermittent support” (score 4 and 3, respectively) is barely needed in a static open-eye balance task of only 10 seconds duration. The under-use of upper scores in *speech* and upper limb subitems indicate that they are biologically rare (in particular in the milder ataxia types), and/or that such advanced patients are no longer seen in ataxia referral centers.

As a third type of limitation, our analysis of longitudinal assessments – with 1-year intervals reflecting the most trial-relevant time interval – revealed non-progression of SARA subitems as major cause of their non-responsive (‘static’) periods. *Speech* was non-responsive due to non-progression from “occasional” (score 3) to “many words difficult to understand” (score 4), or even substantial *improvement* at follow-up (37% and 41% of patients from score 3 and 4, respectively). This may reflect interfering speech therapy^28^, rater bias (e.g., listener experience), and/or effective compensatory mechanisms, especially because assessment of speech in the SARA is based on intelligibility, and on “normal conversation”, which may become verbally scarce and simplified in advanced ataxia. The *speech* subitem of the SARA may benefit from standardized speech tasks with a minimum quantity and complexity of speech production (e.g., reading task or syllable/sentence repetition, as used for outcome assessment in ataxia speech trials^28, 29^). For upper limb subitems of the SARA, we demonstrate fluctuating score decreases (“improvements”) and increases as major cause of their non-progression. We hypothesize that this variability of upper limb subitems is caused by the variable speed-accuracy trade-off that a patient is still free to select despite task instructions (“as fast *and* as precise as possible”, “at moderate speed”), and by difficulties of the rater to visually estimate spatial deviations (dysmetria, tremor amplitude) in an objective and reproducible manner^30^. Subitem *nose-finger* may additionally suffer from an incomplete definition of “kinetic tremor”: it might be interpreted by clinical raters as ‘*rhythmic*, oscillatory movement’ according to movement disorders criteria^31^ (consistent with intention tremor), or as any irregularity due to decomposition and dysmetria of arm movement, as rated in the ICARS^32^.

### Patient-focused functional relevance

Regulatory agencies have expressed concerns not only on the intraindividual and subitem variability of the SARA, but also on the functional relevance of its individual subitems^19^. Our findings from correlation analysis of SARA with FARS-ADL help to inform this discussion. First, the moderate-to-strong correlations of all SARA subitems -except *nose-finger-* with the FARS-ADL indicate that all of these subitems might capture everyday functions in ataxia, given that the FARS-ADL captures complex functional impairment across multiple domains^22^. Second, the discrepancy between poor sensitivity to change and good correlations with FARS-ADL of several SARA subitems indicate that metric problems, not content validity of the respective item, impair responsiveness. This discrepancy was particularly striking for subitem *alternating hand movements*: its strong correlation with FARS-ADL (rho=0.73; comparable to *gait*) suggests that this motor task indeed captures upper limb ataxia functionally meaningful to patients.

### Longitudinal natural history data

Our study provides natural history data for several ARCAs based on the SARA, informing about annualized progression for trial size estimations (mean and variability of slope), and ataxia severities that need to be expected upon trial planning (distribution of intercepts). Specifically, we provide for the first time natural history data for AOA2, SYNE1-ataxia, and AT. Albeit preliminary, given the limited number of observations and heterogeneous time courses, such data are urgently required for trial planning, as treatments are on the horizon for several of these ARCAs -either for the whole ataxia disease type^33^ or for individual patients thereof susceptible to individualized genetic treatments^34^. The progression rates of 1.56 SARA points/year in POLG-ataxia and 0.57 SARA points/year in SPG7 corroborate and extend earlier findings in these ataxias^35, 36^. In ARSACS, where responsiveness of the SARA in annual intervals is controversial^14, 37^, our data suggests that even without ceiling effects, the SARA may not be sensitive to change over 1 year. Larger natural history studies accounting e.g. for different mutations (c.8844delT *SACS* founder mutation vs. other mutations) are needed to address this question.

### Modelling longitudinal progression

As one of the most important findings, our progression and sample size estimations demonstrate that using the SARA as COA in future trials will benefit from specifically considering ataxia severity and subitem composition. Regarding ataxia severity, the total SARA performed best in mild ataxia (baseline SARA: ≤10). This is important as it is this disease stage where disease-modifying therapies might likely be most effective, and which thus represents the disease stratum mainly enrolled in current treatment trials^38^. In moderate ataxia (SARA: 10-25), sensitivity of the SARA was equal to mild ataxia only after stratification for ataxia genotype. This reflects heterogeneity across genetic ataxias at this stage, and indicates that genotype-specific natural history studies may be particularly recommended before applying the SARA as a trial COA to this stage. In advanced ataxia (SARA: >25), a relative 2-3-fold increase in trial size -irrespective of genetic stratification-render the use of the SARA as primary COA in rare genetic ataxias practically impossible. For these patients -almost 15% of our cohort-better COAs, or acceptance of other, non-clinical outcome measures (e.g. fluid^39^ or digital-motor measures^40, 41^) are thus needed.

### Rank-optimized SARA

An improved version of the SARA might be established by a generic ranking and selection of only the most responsive SARA items, across ataxia types (rank-optimized SARA, roSARA). Our study demonstrates that *finger-chase* and *nose-finger* are not beneficial or “neutral” subitems, but in fact detrimental to the performance of the SARA as COA. We show that a data-driven roSARA which omits these two upper limb subitems allows to reduce trial size by up to 25%, and that a composite of only 3-4 most responsive subitems may appear superior to the total SARA, but still have suboptimal sensitivity. This finding extends recent findings limited to SCA3^15^ to a large across-genotype cohort, thus demonstrating their general applicability to genetic ataxias. In combination with the suggested metric optimizations, such a rank-optimized SARA might help to facilitate its regulatory qualification for upcoming trials^42^.

### Limitations of the study

In line with the concept and development of the SARA^1^, our across-genotype analyses assume a unified core construct of *ataxia* underlying the large overall data aggregate. Although we controlled for genotype in the common ARCA cohort, key findings which could not be analyzed on genotype-specific level (e.g. the added value of the roSARA) would warrant further validation in larger cohort studies of the respective genetic ataxia type. However, our study is fully in line with smaller genotype-specific studies, e.g. in FA^26^ or SCA3^15^, which focused on some of the aspects now identified here as part of a larger set of findings with general applicability across genetic ataxias. Moreover, for many of these ultra-rare ataxias, genotype-specific validation will not be realistic in the near future. Taking ARCAs as whole group allows to delineate a distribution range of possible disease trajectories across all ARCAs, including the rarest ones. This can be used for estimating and narrowing the probability distributions of disease trajectories even for ataxias where no within-genotype natural history studies can be done, using it e.g. as prior for Bayesian trajectory estimations^43^. This study is also limited by the fact that it resorts to multicenter registry rather than clinical trial data. While this adds data variability, such use of real-world data allows to indeed apply the findings reported here to real-world applications of SARA as COA^44^. Finally, our sample size calculations aimed to compare the *relative* sensitivity of the SARA between ataxia severities and SARA composites as a generic, i.e. across-genotype COA for ataxia. While this allows to illustrate the COA properties of the SARA for ataxia as a core construct across the multitude of genetic ataxias, the heterogeneity of underlying ataxia types is expected to increase sample sizes, and extrapolation to genotype-specific treatment trials has to be interpreted with caution. However, in the absence of treatment trial data for literally all ataxias analyzed here (except FA), our sample size calculations provide at least a first starting point allowing to estimate relative sample size magnitudes for upcoming trial planning. As more data become available in rigorously performed trial-like natural history studies (e.g., ARSACS, SPG7, or RFC1)^45, 46^ and future treatment trials, this will allow to validate and further refine the findings of the present study in disease-specific trial contexts.

## Data Availability

All data produced in the present study are available upon reasonable request to the authors.

